# The newly introduced SARS-CoV-2 variant A222V is rapidly spreading in Lazio region, Italy

**DOI:** 10.1101/2020.11.28.20237016

**Authors:** Barbara Bartolini, Martina Rueca, Cesare Ernesto Maria Gruber, Francesco Messina, Emanuela Giombini, Giuseppe Ippolito, Maria Rosaria Capobianchi, Antonino Di Caro

## Abstract

A new SARS-CoV-2 clade (GV) characterized by S substitution A222V, first reported from Spain in March, is rapidly spreading across Europe. To establish the A222V variant involvement in the infection rise in Italy, all GISAID sequences from Italy and those from our Laboratory (Lazio) in the period June-October were analysed. A222V, first recognized in August, represents 11.2% of sequences in this period, reaching 100% of autochthonous sequences in October, supporting increased GV circulation in Italy.

## Background

Monitoring the emergence and spread of viral variants is a critical issue of virological surveillance, particularly in the present SARS-CoV-2 pandemic. In fact, it is essential to study the epidemiological dynamic and for the prompt recognition of mutations that may i. affect diagnostic recognition, ii. hamper the effectiveness of vaccines and iii. reduce the effects of therapeutic intervention [1,2]. A new variant carrying the A222V substitution in the spike protein (S) was first observed in Spain in March 2020, and rapidly spread since June across Europe [3], seeding a new clade (GV), that is at present accounting for about 6.6% of all GISAID submitted genomes (https://www.gisaid.org/, accessed Nov 3, 2020). Until recently, only one GV sequence was reported to GISAID from Italy, occurring in Sardinia, collected in August 25. We regularly perform whole genome sequence analysis to support the surveillance service in Lazio region, Italy. To investigate if this new variant plays a role in the abrupt rise of infections that is currently affecting our country, we describe the sequences obtained from samples collected in our Laboratory (Rome, Lazio), covering the period of June-October.

## Methods and main results

A total of 57 whole genome sequences obtained in our laboratory from June 1st and October 31st were included in the analysis. More in details, we included sequences obtained for surveillance purpose in the period June-July (n=46), sequences from persons with known travel history, sampled in the second half of August, when the resurgence of the outbreak in Italy started (n=5) and sequences from the third week of October, to represent the presently circulating virus variants (n=6).

Whole genome sequencing (WGS) was performed on residual respiratory samples by using Ion AmpliSeq™ SARS-CoV-2 research panel on GeneStudio™ S5 Prime System (Thermofisher). Raw reads with mean quality Phred score >20 were selected and trimmed using Trimmomatic software v.0.36 [4]. SARS-CoV-2 genomes were assembled using reference-based assembly method, with BWA v.0.7.12 [5] and Samtools v.1.3.1 [6]. Assembled contigs were then verified using Geneious 2019.2.3.

Besides the sequences obtained on our laboratory, we also included all the sequences of SARS-CoV-2 strains collected from Italy between June 1^st^ and October 31^st^ and retrieved from GISAID [7], resulting in a total of 89 good quality sequences. These were clustered at 100% using CD-HIT v.4.6 software [8], providing 57 unique sequences, that were included in the phylogenetic three. Maximum likelyhood phylogenetic analysis was performed with IQ-TREE v.1.6.12, using General Time Reversible with empirical base frequencies plus Gamma model (GTR+F+I) and 1000 replicates; Wuhan-Hu-1 strain was adopted as phylogenetic outgroup (NCBI accession number: MN908947.3).

Samples collection date, sample type, viral clade and travel history of patients, are reported in table 1.A phylogenetic tree (Figure 1) encompassing all the 57 unique SARS-CoV-2 sequences from Italy, collected from June 1^st^, 2020 was built. As can be seen, the phylogenetic tree clearly shows the presence of one cluster of sequences epidemiologically correlated, belonging to the clade GR and occurred in June in Rome, Lazio, that will be described in details elsewhere. Nine sequences obtained in our laboratory, and one reported to GISAID from Sardinia by Laboratorio Biologia Molecolare SarsCov2 Ospedale S. Francesco ATS-ASSL Nuoro (SAR-ATS-246, August 25), harboured the A222V substitution and are included in the GV clade, accounting for 17.5% of contemporary Italian sequences included in the phylogenetic tree. The study subjects harbouring this viral strain were returning travellers in the first period (from Sardinia: INMI 85, August; Spain: INMI82 and INMI84, August), and patients without known travel history later on (i.e. in October). In the period June-October other clades, namely GR, G and GH, co-circulated in Italy, accounting for 68.4%, 8.8% and 5.3% of the contemporary Italian sequences included in the phylogenetic tree, respectively. In particular, other travel related isolates collected at INMI in August (INMI81 from Croatia and INMI83 from Malta) were included in clades GR and GH, respectively. Further, the sequences of three isolates collected in October in Campania and reported in GISAID (EPI_ISL_602304, 584071 and 584069) from U.O. Diagnostica Virologica Dip. Sanità Animale Istituto Zooprofilattico del Mezzogiorno, Portici, Napoli, belong to clade G.

**Table 1.**
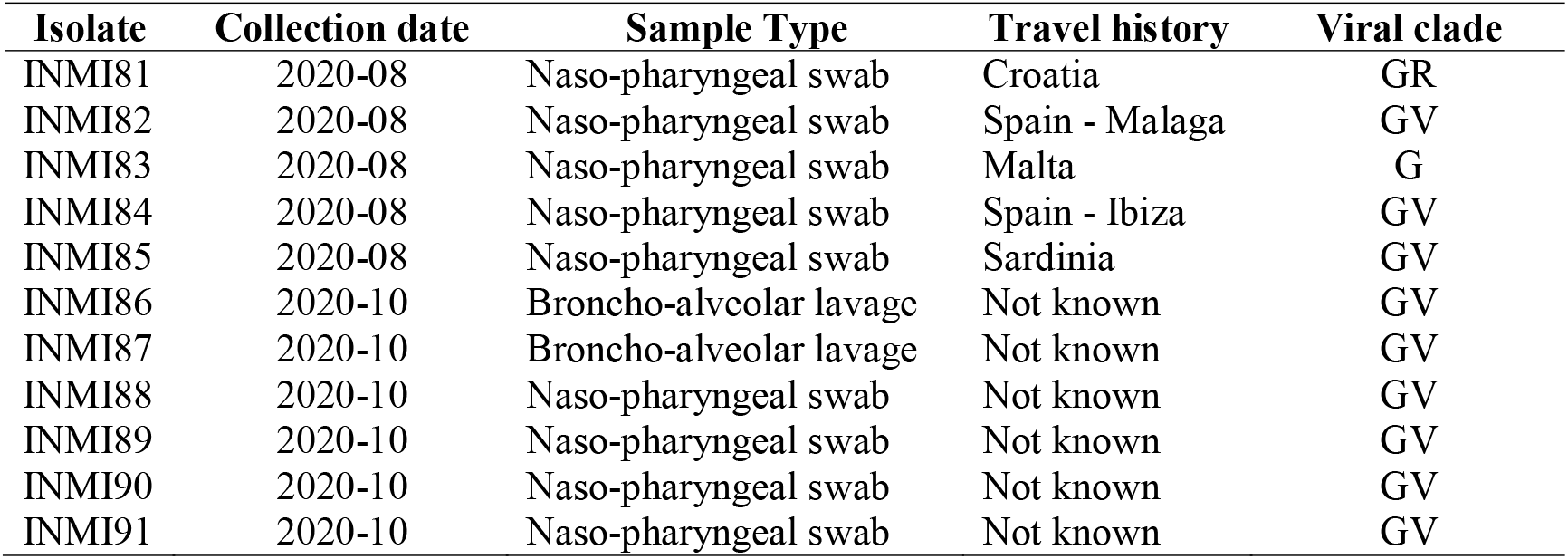
Samples collection date, sample type, viral clade and travel history of study patients

**Figure 1:**
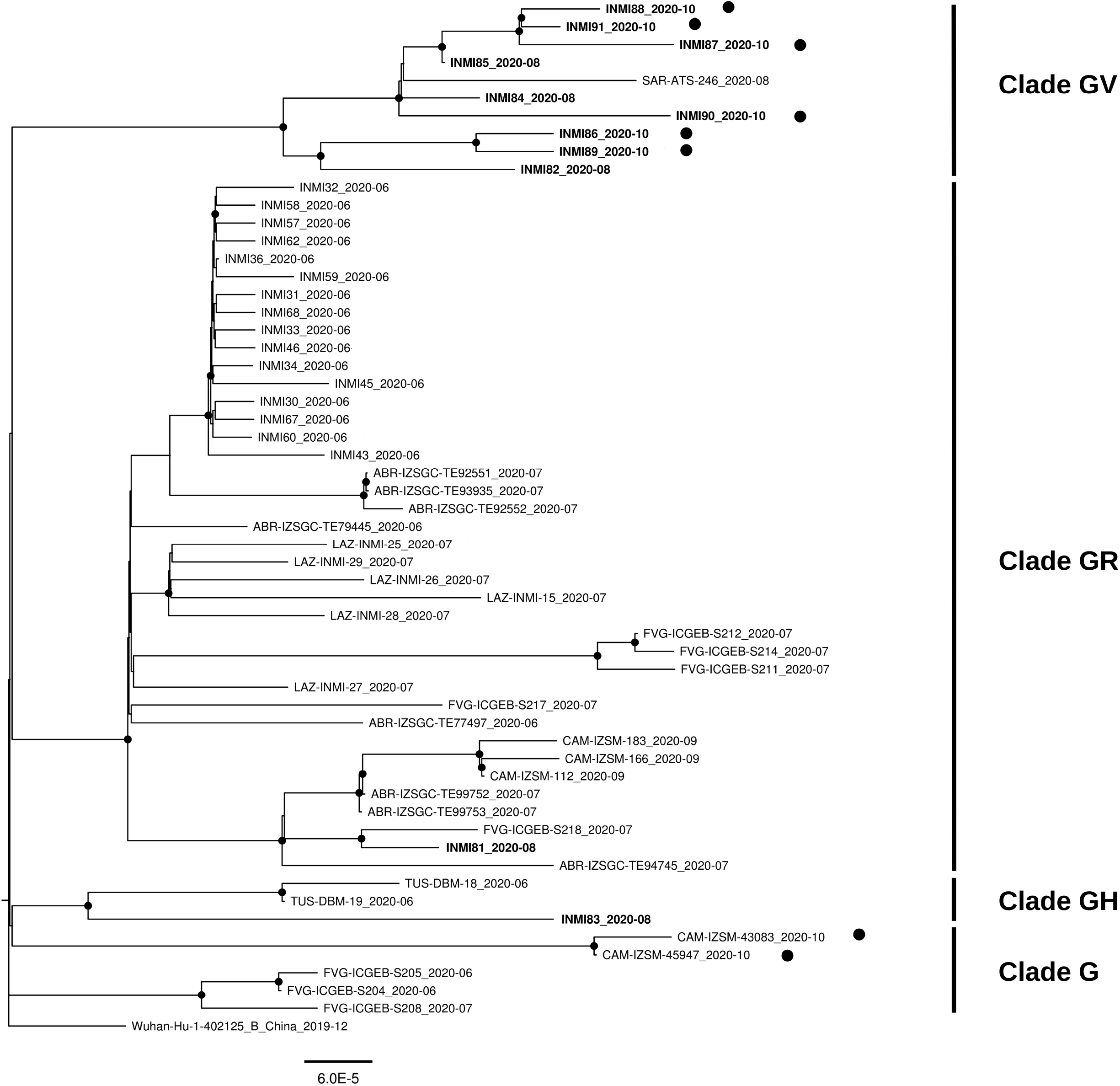
Maximum likelihood phylogenetic tree of all SASR-CoV-2 unique sequences obtained in Italy in from 1^st^ June and 2^nd^ November 2020 (n=57), including 33 sequences obtained in our laboratory, at the National Institute for Infectious Diseases (INMI) in Rome, Italy, and 24 sequences from other Institutions in Italy. Sequences obtained at INMI are indicated by the name that also includes the collection date. To underline the chronological order of sequences obtained in our laboratory, sequences obtained from August onward are highlighted in bold. Italian sequences obtained in late October are highlighted with a bullet. Clades are indicated on the side of the tree, following the GISAID nomenclature. All nodes with bootstrap values higher than 75 are highlighted with a black point.

## Discussion

Despite the small number of sequences so far reported from Italy, our data support a suddenly increased circulation of the GV clade in Italy. The A222V variant represents 11.2% of sequences in the period June-October 2020. In particular, the GV infections initially observed in Italy, occurring in August, were observed in Sardinia and travellers returning to Lazio region from Sardinia and Spain. On the contrary, all the GV sequences obtained in October were from patients without travel history, therefore presumably autochthonous. Taken together, these data suggest the introduction of clade GV in Italy in August, possibly in Sardinia and in other regions (including Lazio) by returning travellers, followed by extensive local spread that is still ongoing. It is not known at the moment whether other Italian regions are experiencing rising circulation of this new clade, and more data are necessary to better describe the virus dynamics and spread in our country.

Although A222V mutation is located in a region of spike protein distant from the cell receptor binding site [3], an advantage in spreading efficiency cannot be excluded and needs further investigation, also in view of its location in a putative B cell epitope [9] that may affect immune recognition.

## Data Availability

Sequence data have been posted on GISAID www.gisaid.org

## Ethical approval

This work was performed within the framework of the COVID-19 outbreak response and surveillance program, and has been approved by the INMI Ethical Committee (“Comitato Etico INMI Lazzaro Spallanzani IRCCS/Comitato Etico Unico Nazionale Covid-19”; issue n. 214/20-11-2020).

## Acknowledgments

We gratefully acknowledge the contributors of genome sequences of the newly emerging coronavirus, i.e. the Originating and Submitting Laboratories, for sharing their sequences and other metadata through the GISAID Initiative, on which this research is based.

## Disclaimer

The sequences have been deposited in GISAID with accession IDs:

EPI_ISL_609990, EPI_ISL_609989, EPI_ISL_609999, EPI_ISL_609998, EPI_ISL_609997, EPI_ISL_609996, EPI_ISL_609995, EPI_ISL_609994, EPI_ISL_609993, EPI_ISL_609992, EPI_ISL_609991

